# Decreased brain pH correlated with progression of Alzheimer’s disease neuropathology: a systematic review and meta-analyses of postmortem studies

**DOI:** 10.1101/2024.04.30.24306603

**Authors:** Hideo Hagihara, Tsuyoshi Miyakawa

**Author notes:** Correspondence: Hideo Hagihara, Division of Systems Medical Science, Center for Medical Science, Fujita Health University, 1-98 Dengakugakubo, Kutsukake-cho, Toyoake, Aichi 470-1192, Japan., ORCID ID: 0000-0001-9602-9518, Tsuyoshi Miyakawa, Division of Systems Medical Science, Center for Medical Science, Fujita Health University, 1-98 Dengakugakubo, Kutsukake-cho, Toyoake, Aichi 470-1192, Japan., ORCID ID: 0000-0003-0137-8200.

## Abstract

**Background:** Altered brain energy metabolism is implicated in Alzheimer’s disease (AD). Limited and conflicting studies on brain pH changes, indicative of metabolic alterations associated with neural activity, warrant a comprehensive investigation into their relevance in this neurodegenerative condition. Furthermore, the relationship between these pH changes and established AD neuropathological evaluations, such as Braak staging, remains unexplored.

**Methods:** We conducted quantitative meta-analyses on postmortem brain and cerebrospinal fluid pH in patients with AD and non-AD controls, using publicly available demographic data. We collected raw pH data from studies in the NCBI GEO, PubMed, and Google Scholar databases.

**Results:** Our analysis of 17 datasets (457 patients and 315 controls) using a random-effects model showed a significant decrease in brain and cerebrospinal fluid pH in patients compared to controls (Hedges’ *g* = –0.54, *p* < 0.0001). This decrease remained significant after considering postmortem interval, age at death, and sex. Notably, pH levels were negatively correlated with Braak stage, indicated by the random-effects model of correlation coefficients from 15 datasets (292 patients and 159 controls) (adjusted *r* = –0.26, *p* < 0.0001). Furthermore, brain pH enhanced the discriminative power of the *APOE*ε4 allele, the most prevalent risk gene for AD, in distinguishing patients from controls in a meta-analysis of four combined datasets (95 patients and 87 controls).

**Conclusions:** The significant decrease in brain pH in AD underlines its potential role in disease progression and diagnosis. This decrease, potentially reflecting neural hyperexcitation, could enhance our understanding of neurodegenerative pathology and aid in developing diagnostic strategies.

## Introduction

Alzheimer’s disease (AD) is a prevalent neurodegenerative disease worldwide, posing one of the most significant health burdens of the 21^st^ century (Prince et al., 2015; Scheltens et al., 2016). Genetic studies have demonstrated that AD has a high heritability, with multiple genetic factors contributing to the disease. Over 50 risk loci have been associated with AD at a genome-wide significance level (Sims et al., 2020). Among them, point mutations leading to the ε4 allele of apolipoprotein E (*APOE*) are known to be the most potent genetic risk factor for the common sporadic and late-onset forms of AD, as well as influencing rarer familial and early-onset forms of the disease (Saunders et al., 1993; Belloy et al., 2019; Sims et al., 2020; Serrano-Pozo et al., 2021). While the *APOE* gene is suggested to have no association with disease progression (Wilkosz et al., 2010), many other factors can influence the development and progression of pathophysiological changes in the brains of individuals with AD. In particular, pH in the brain could be significant in the pathophysiology of AD, as low pH has been reported to exacerbate the aggregation of the amyloid-β peptide and hyperphosphorylation of Tau protein, key pathological hallmarks of the disease (Atwood et al., 1998; Basurto-Islas et al., 2013; Decker et al., 2021). To date, studies of pH changes in the brain of AD have been limited and the results are inconsistent. One postmortem study involving four available datasets has reported a decrease in brain and cerebrospinal fluid (CSF) pH in patients with AD compared to control subjects in each dataset (Decker et al., 2021). The results of the magnetic resonance spectroscopy (MRS) studies were inconsistent: a decrease (Lyros et al., 2020), no significant change (Mandal et al., 2012), and an increase (Mecheri et al., 1997; Rijpma et al., 2018) in pH have been observed in the hippocampus and other regions of the brain of patients with AD. Furthermore, no study has yet elucidated the relationship between brain pH and the progression of AD, particularly in the context of Braak staging – a well-established neuropathological method for AD diagnosis based on neurofibrillary tangles (Braak and Braak, 1991).

Decreased brain pH has been consistently suggested in schizophrenia and bipolar disorder by meta-analysis of MRS and postmortem studies (Dogan et al., 2018; Hagihara et al., 2018; Pruett and Meador-Woodruff, 2020). We recently found similar phenomenon in major depressive disorder in a systematic review and meta-analysis of postmortem studies (Hagihara and Miyakawa, submitted). A decrease in brain pH is suggested to be associated with an increase in lactate level under these psychiatric disorders (Prabakaran et al., 2004; Stork and Renshaw, 2005; Halim et al., 2008), which could be due to metabolic changes resulting from mitochondrial dysfunction and/or increased glycolysis due to neuronal hyperexcitation. An increase in brain (Mullins et al., 2018; Hirata et al., 2024) and CSF (Liguori et al., 2015, 2016) lactate levels has been observed in patients with AD compared to control subjects, suggesting common metabolic alterations and neuronal hyperexcitability across such psychiatric and neurodegenerative disorders.

In view of the limited studies and contradictory results regarding pH changes in AD, in this study we conducted a systematic review and quantitative meta-analyses of pH in the postmortem brain and CSF in patients with AD compared to non-AD control subjects, utilizing publicly available demographic data. Furthermore, we conducted meta-analyses to investigate the association between pH and disease progression assessed by the Braak staging and the discriminative ability of brain pH in distinguishing patients with AD from control subjects.

## Methods

### Ethics approval and consent to participate

This study was performed based on publicly available data and no separate ethical approval was required.

### Identification and selection of eligible datasets of postmortem brain and CSF pH

Datasets were selected based on the availability of raw pH data of individual subjects. We searched the National Center for Biotechnology Information (NCBI) Gene Expression Omnibus database (GEO), PubMed, and Google Scholar for studies reporting individual pH data with key words “Alzheimer’s disease”, “brain pH”, “postmortem” and “Braak”. The dataset searches followed the Preferred Reporting Items for Systematic Reviews and Meta-Analyses guidelines (Page et al., 2021) and were conducted on October 2023. The workflow of the selection of pH datasets is shown in Fig. 1. Articles were screened first based on their titles, followed by a subsequent evaluation of their abstracts to determine eligibility for full-text review. For records identified in PubMed and Google Scholar, inclusion criteria were of studies involving human AD versus non-AD control comparisons (CON) using postmortem brain or CSF samples and the availability of full text and individual raw data. To minimize the inclusion of overlapping samples across different studies, we attempted to identify and remove duplicated samples from studies that used partially overlapping samples from the same source, and then combined the remaining data into a single dataset. If conditions other than CON or AD were present within the study, they were excluded from the current analysis. We also obtained the accompanying demographic information from the identified datasets (i.e., postmortem interval (PMI), age at death, sex, Braak stage, and *APOE* genotype). In the dataset of Ueberham 2006, Braak stage labeled as V−VI was treated as 5.5. In the datasets that provided ABC score, scores 1, 2, and 3 in category “B” (representing Braak stage) were treated as stages 1.5, 3.5, and 5.5, respectively, as score 1 encompasses Braak stage I or II, score 2 corresponds to stage III or IV, and score 3 to stages V or VI (Montine et al., 2012). A score of 0 indicates the absence of neurofibrillary pathology. In the dataset of Gabitto 2023, age labeled as 90+ was considered as 90.

**Figure 1.**
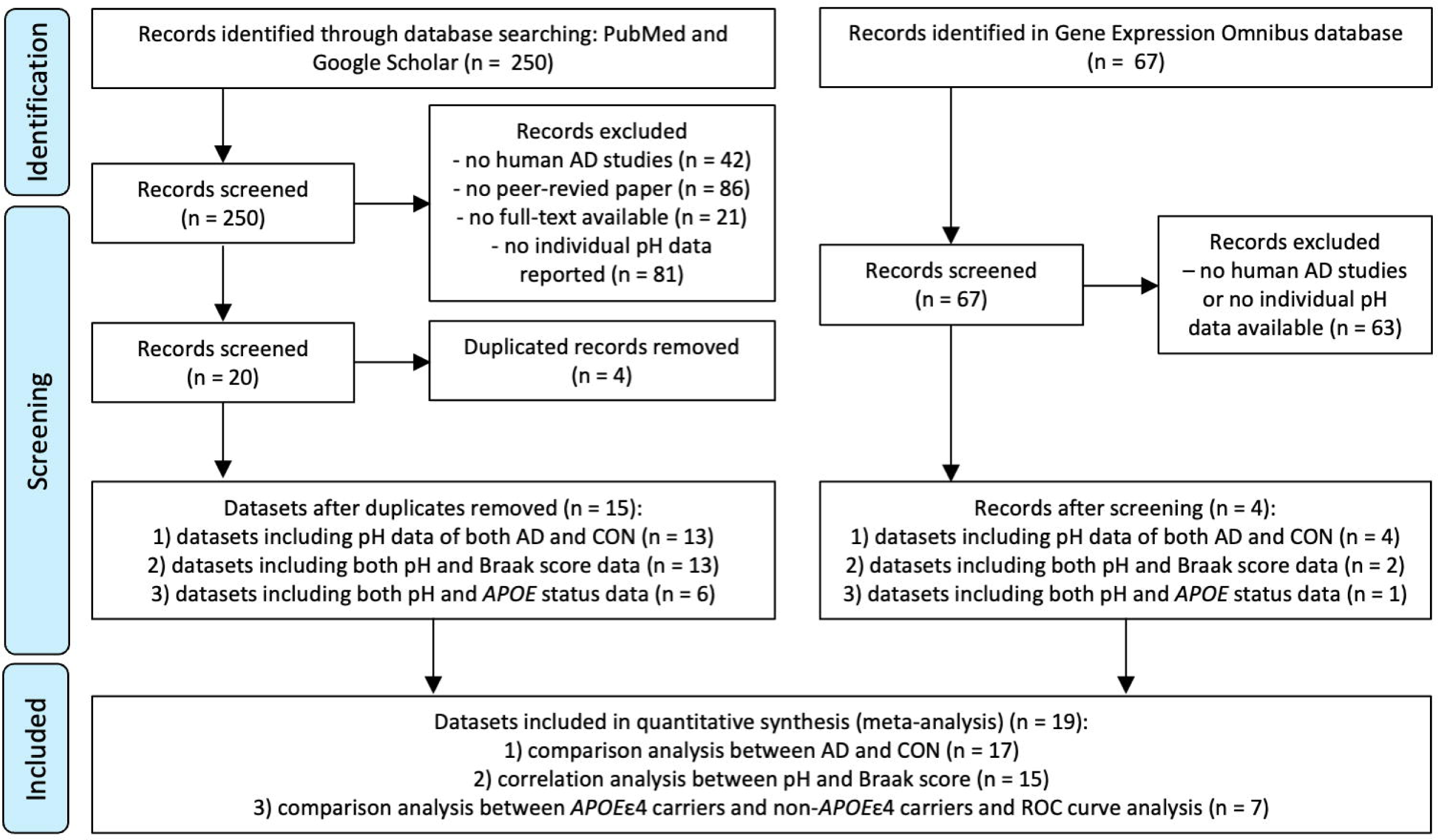
Workflow of the selection of postmortem brain and CSF pH datasets.

## Data analysis

Effect size, measured by Hedges’ *g*, and standard error was calculated within each dataset, and the random effects model was applied using the function metagen in the R package meta (version 6.5-0). Analysis of covariance (ANCOVA) were performed using the GLM procedure in SAS (version 3.81; SAS Institute Inc., Cary, NC, USA). For a correlation analysis, we calculated the Pearson correlation coefficient within each dataset. To estimate the standard error of the correlation coefficient, we used the formula: SE*_r_* = (1−*r*^2^)/(*n*−2)^1/2^, where *r* is the Pearson correlation coefficient, and *n* is the number of samples in the correlation. We then applied the random effects model to the correlation coefficient (*r*) and its standard error (SE*_r_*), as described above. Z-score transformation, a method of data normalization for direct comparison between different conditions, was applied to each pH value using individual subject data within each dataset according to the following formula: Z-score = (value_P_ – mean value_P1…Pn_)/standard deviation_P1…Pn_, where P is any pH or lactate and P1…Pn represent the aggregate measure of all pH values. We conducted the receiver operating characteristics (ROC) curve analysis to evaluate the discriminative ability of the *APOE*ε4 status, pH level, and their combination in identifying patients with AD from control subjects, as assessed by the numerical gains in the area under the curve (AUC). The ROC analyses were performed using GraphPad Prism 8 (version 8.4.2; GraphPad Software, San Diego, CA). For ROC analyses, z-score transformation, a conventional method for data normalization to enable direct comparison between different conditions, was applied to pH values within each dataset, given their heterogeneity across datasets. Multiple logistic regression analyses were applied to the z-score of pH level, *APOE*ε4 status (yes = 1, i.e. ε3/ε4 and ε4/ε4; no = 0), and their combination, respectively.

## Results

### Search results

Following screening the 250 and 67 records retrieved from the searches in PubMed and Google Scholar, and the NCBI GEO database, 20 and 4 studies were identified to report raw pH data, respectively (Fig. 1). After removing duplicates, a total of 19 datasets were processed for the meta-analysis: 1) 17 datasets for comparison analysis of pH between patients with AD and control subjects (Fig. 2); 2) 15 for correlation analysis between pH and Braak stage (Fig. 3); and 3) seven for comparison analysis of pH between *APOE*ε4 carriers and non-*APOE*ε4 carriers (Supplementary Fig. 3) and ROC curve analysis (Fig. 4). A summary of the datasets, which contains information on pH, PMI, age at death, and sex, is presented in Supplementary Table 1. The source and references of the datasets used are also included in Supplementary Table 1. The raw data analyzed in this study are provided in Supplementary Table 2.

**Figure 2.**
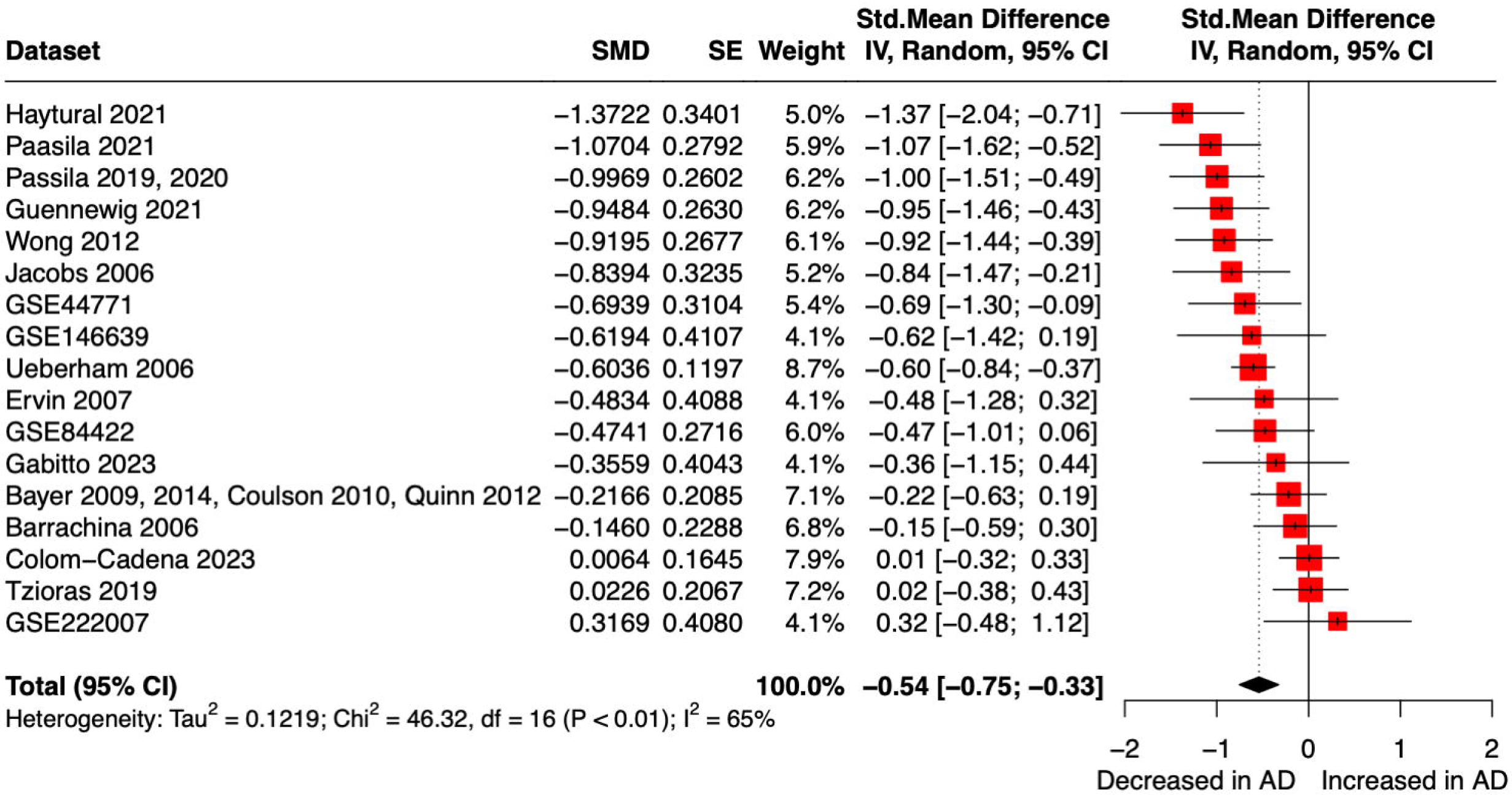
Decreased pH in the postmortem brain and CSF of patients with AD. Forest plot of meta-analysis comparing postmortem brain and CSF pH between patients with AD and non-AD control subjects. 95% CI, 95% confidence interval; SE, standard error; SMD, standardized mean difference.

**Figure 3.**
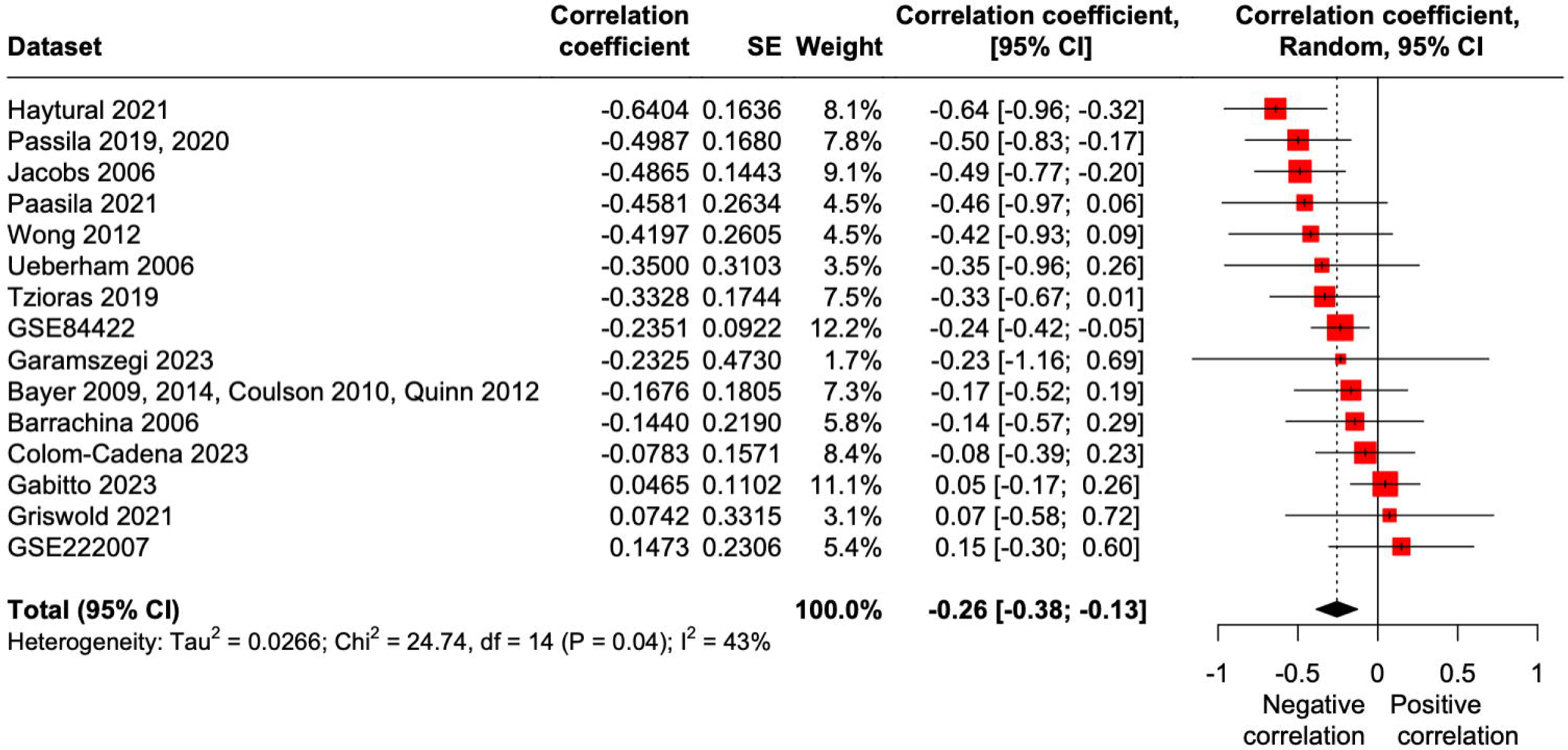
Negative correlation between brain pH and Braak stages. Forest plot of meta-analysis of correlation between brain pH and Braak stages. 95% CI, 95% confidence interval; CC, correlation coefficient; SE, standard error.

**Figure 4.**
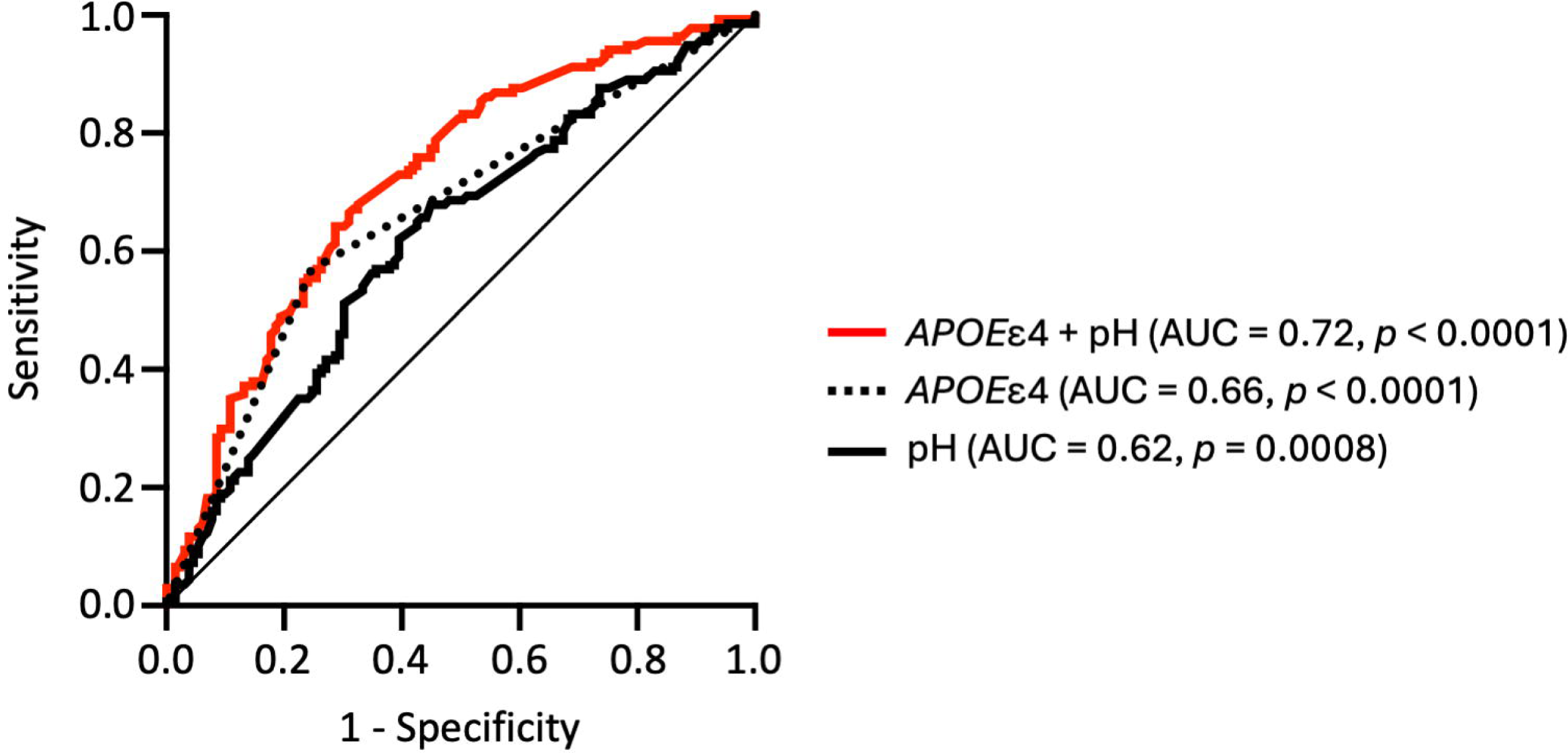
Brain pH level increases discriminative accuracy of *APOE*_ε_4 status for AD. ROC curves for brain pH level, *APOE*ε4 status, and their combination to assess the accuracy of prediction of AD against controls. AUC, area under the curve.

### Decreased brain and CSF pH in AD

In the 17 datasets obtained (Supplementary Table 1), six showed a significant decrease in pH in patients with AD compared to the corresponding controls within each dataset (GSE44771, *p* = 3.51 × 10^-7^; GSE84422, *p* = 0.044; Guennewig 2021, *p* = 0.0016; Haytural 2021, *p* = 0.015; Jacobs 2006, *p* = 0.021; Passila 2019, 2020, *p* = 0.033; two-tailed unpaired t-test). The others showed no significant difference in pH between patients and controls. No datasets showed a significant increase in pH. Funnel plot and Egger’s test showed no evidence of publication bias for the 17 datasets (*t* = –0.89, *p* = 0.39; Supplementary Fig. 1). The meta-analysis using the random effects model revealed a significant decrease in pH in patients with AD compared control subjects (Hedges’ *g* = –0.54, *z* = –4.97, *p* < 0.0001, 95% CI = [-0.75; –0.33]) (Fig. 2). The *I^2^* test showed high heterogeneity suggesting significant differences between datasets (*I^2^* = 65%, *p* < 0.01). In a two-way ANCOVA, with the dataset and diagnosis as the main factors, the effect of diagnosis remained significant (mean square (MS) = 2.25, *F* = 23.78, *p* < 0.0001), while no significant effects were observed for PMI (MS = 0.12, *F* = 1.24, *p* = 0.26), age at death (MS = 0.35, *F* = 3.71, *p* = 0.055), or sex (MS = 0.29, *F* = 3.06, *p* = 0.081). The effect of dataset was significant (MS = 1.31, *F* = 13.85, *p* < 0.0001). These results suggest that, while the pH varied between datasets, it was decreased in patients with AD compared to controls, independently of the covariates examined.

### A decrease in pH is associated with progressive brain pathological changes

The current assessment of the severity of pathological changes in AD is mainly based on the Braak staging, where stages I–II represent clinically silent cases, stages III–IV incipient AD, and stages V–VI fully developed AD (Braak and Braak, 1991). We then examined the relationship between brain pH levels and the Braak stage. In the 15 datasets obtained (Supplementary Table 1), four showed a significant negative correlation between pH levels and Braak stage in each dataset (GSE84422, *r* = –0.24, *p* = 0.015; Passila 2019, 2020, *r* = –0.50, *p* = 0.018; Jacobs 2006, *r* = –0.49, *p* = 0.0064; Haytural 2021, *r* = –0.64, *p* = 0.010). The others showed no significant correlation within datasets. The meta-analysis, employing a random effects model, revealed a significant negative correlation between pH levels and Braak stage (adjusted *r* = –0.26, *p* < 0.0001, 95% CI = [-0.38; –0.13]) (Fig. 3). This negative correlation was corroborated by linear regression analysis on 15 combined datasets, standardized using z-score normalization, which showed a modest but significant relationship (*r* = –0.20, *p* = 1.37 × 10^-5^; Supplementary Fig. 2). These results suggest that lower pH levels are associated with higher severity of the disease.

The brain pH also showed significant negative correlations with other pathological and clinical rating scores, including clinical dementia rating (*r* = –0.22, *p* = 0.026), average neuritic plaque density (*r* = –0.25, *p* = 0.0095), sum of Consortium to Establish a Registry for Alzheimer’s Disease (CERAD) rating scores in multiple brain regions (*r* = –0.34, *p* = 0.0004), and sum of neurofibrillary tangles density in multiple brain regions (*r* = –0.24, *p* = 0.014) (Supplementary Fig. 3). These results support the idea that lower pH levels are associated with higher severity of the disease.

### Brain pH improves discriminative ability of *APOE***_ε_**4 status in AD

The ε4 allele of apolipoprotein E (*APOE*) is the strongest genetic risk factor for sporadic AD (Belloy et al., 2019). While *APOE*ε4 status did not affect brain pH level either in patients with AD or control subjects (Supplementary Fig. 4), it exhibited a significant discriminative ability in identifying patients from controls, as determined by the ROC curve analysis using seven datasets consisting of 264 subjects (Fig. 4, Supplementary Table 1). The ROC AUC values of the single-predictor model were 0.66 for *APOE*ε4 status (95% CI: 0.60–0.73, *p* < 0.0001) and 0.62 for the pH level (95% CI: 0.55–0.69, *p* = 0.0008). When the pH level was added to the *APOE*ε4 status, the ROC AUC value increased to 0.72 (95% CI: 0.66–0.78, *p* < 0.0001; *p* for difference 0.0075) (Fig. 4). The discriminative accuracy of *APOE*ε4 status was improved by the combination with the pH level, suggesting that these two variables have a complementary value.

## Discussion

The present study suggested that brain pH is decreased in patients with AD compared to control subjects even when PMI, age, and sex were considered as potential confounding factors. We recently showed that a decrease in brain pH in AD can be estimated through the analysis of gene expression patterns (Hagihara et al., 2023), which aligns with the findings of this study. Furthermore, the increased levels of brain lactate in patients with AD provide additional support for the observed decrease in pH.

A decrease in brain pH has been linked to an increase in lactate levels in schizophrenia and bipolar disorder (Prabakaran et al., 2004; Stork and Renshaw, 2005; Halim et al., 2008). An increase in lactate levels is considered indicative of metabolic alterations resulting from mitochondrial dysfunction and/or increased glycolysis due to neuronal hyperexcitation. Lactate is thought to be a relatively strong acid due to its ability to almost completely dissociate into H^+^ ions and lactate anions at cellular pH (Siesjö, 1985). Indeed, hyperexcitation in certain neurons and/or neural circuits has been suggested in the etiology of AD (Busche and Konnerth, 2016; Murano et al., 2019; Maestú et al., 2021). Furthermore, an increase in brain (Mullins et al., 2018; Hirata et al., 2024) and CSF (Liguori et al., 2015, 2016) lactate levels has been observed in patients with AD compared to control subjects. Neuronal hyperexcitation (Palop et al., 2007; You et al., 2017) and increased brain lactate and decreased brain pH levels (Hagihara et al., 2024) have been observed in a well-known mouse model of AD harboring the human amyloid precursor protein (APP) mutant allele that is linked to familial AD. At a cellular level, glycolysis and subsequent lactate release are stimulated by the uptake of the neurotransmitter glutamate in astrocytes following neuronal activation, as demonstrated in an *in vitro* study (Pellerin and Magistretti, 1994). Astrocytes are considered to prefer lactate production in the brain. A shift towards excitation would increase energy demands in neurons, potentially prompting astrocytes to elevate lactate production, which in turn could lower brain pH. Indeed, increased lactate levels, accompanied by astrocyte activation, have been observed in the brains of individuals of AD (Hirata et al., 2024). Therefore, the observed decrease in brain pH may be attributed to elevated lactate levels induced by neuronal hyperexcitation in AD. Activity-dependent acidosis has been suggested to suppress neuronal activity in animal models of neonatal febrile seizures and is proposed as an intrinsic mechanism for the self-termination of seizures (Chesler and Kaila, 1992; Schuchmann et al., 2006; Pavlov et al., 2013). Further research is needed to determine whether a decrease in brain pH is related to such compensatory mechanisms in chronic degenerative diseases.

We found a negative correlation between brain pH and Braak stage, suggesting a lower brain pH as the disease course progresses. Braak staging of AD is based on the propagation of tau neurofibrillary tangles in the brain and the stage of tau pathology is associated with cognitive impairment (Braak and Braak, 1991). It is hypothesized that soluble amyloid-β oligomers cause hyperphosphorylation of tau and the accumulation of neurofibrillary tangle (Zheng et al., 2002); however, the local relationship between amyloid and tau was restricted in several brain regions (Iaccarino et al., 2018), suggesting additional causes for tau accumulation. Interestingly, neuronal hyperexcitation has been suggested to enhances tau protein translation in neurons, which may lead to the pathological accumulation of tau in soma and dendrite of neuronal cells (Kobayashi et al., 2017, 2019). Therefore, a decrease in pH and the accumulation of tau may occur in parallel due to neuronal hyperexcitation. Furthermore, acidic conditions could increase the abnormal hyperphosphorylation of tau protein *in vitro* (Basurto-Islas et al., 2013). These potential direct and indirect relationships between pH and tau pathology could contribute to the observed negative correlation between pH and Braak stage. Brain pH may serve as a surrogate for Braak stage.

We found that brain pH significantly improved discriminative accuracy of *APOE*ε4 status in distinguishing patients with AD from control subjects. A previous *in vivo* MRS study showed that brain pH improved diagnostic accuracy of brain metabolite (i.e., N-acetylaspartate/creatine ratio) for AD (Lyros et al., 2020). While brain pH alone exhibited relatively low (AUC of 0.62 in this study) to moderate (AUC of 0.81 in (Lyros et al., 2020)) accuracy for distinguishing patients with AD from control subject, combining it – measured in CSF or by MRS – with other markers could be a valuable biological strategy to assist in the diagnosis of AD.

Regarding potential bias in this study, the Egger’s regression test on the funnel plot showed no significant publication bias across datasets analyzed. Additionally, relying on demographic rather than primary outcome data may minimize typical outcome bias.

A significant limitation of this study may involve the lack of consideration for other potential confounding factors affecting postmortem brain pH than age at death, PMI, and sex. Factors prior to death, or agonal states, and the cause and manner of death can influence postmortem brain pH through alterations in blood oxygenation, brain perfusion, and biochemical content (Lewis, 2002; Tomita et al., 2004). If patients with AD experienced prolonged agonal conditions compared to controls in the datasets used, we cannot rule out the possibility that the observed decrease in pH was influenced by such factors. It is technically difficult to exclude the influence of potential confounding factors in human studies. In this circumstance, studies in animal models can be useful alternatives to confirm whether changes in brain pH are involved in the pathophysiology of neuropsychiatric disorders, as they are essentially exempt from such confounding factors. In our recent study, we showed that APP transgenic AD model mice exhibited a decrease in brain pH compared to corresponding control mice (Hagihara et al., 2024), supporting the notion that this phenomenon is intrinsically related to pathophysiology of AD rather than mere an artifact.

In conclusion, this study suggested that postmortem brain and CSF pH is decreased in patients with AD compared to control subjects. The decrease in pH may occur independent of having *APOE*ε4 genotype, the most prominent genetic risk of AD. Neuronal hyperexcitation may lead to the decrease in pH, which may be exacerbated in the disease course. It remains undetermined whether such a decrease in pH is associated with either beneficial or detrimental effects on psychiatric conditions, which is required to be addressed in the future studies.

## Acknowledgements

The authors thank Yoko Kagami and Harumi Mitsuya for their administrative assistance.

## Data availability

The raw data analyzed in this study are provided in Supplementary Table 2.

## Author contributions

HH and TM conceived this study. HH performed data search, analyzed data, and drafted the manuscript. All authors contributed to revising the manuscript and approved its final version.

## Additional information

The review was not registered elsewhere. The review protocol was not prepared.

## Supplementary information

**Supplementary Table 1. Summary of datasets used in this study**

**Supplementary Table 2. Raw data analyzed in this study**

**Supplementary Figure 1. Funnel plot of the included datasets.** Each plot represents a dataset. SE, standard error; SMD, standardized mean difference.

**Supplementary Figure 2. Scatter plot depicting the negative correlation between pH (z-score) and Braak stages.** A z-score was calculated for each subject within the dataset to standardize pH measurements. Each dot represents the data for an individual subject. *r*, Pearson correlation coefficient.

**Supplementary Figure 3. Significant correlation between a decrease in brain pH and an increase in disease severity.** Data of GSE84422 was analyzed. Scatter plot of correlation between brain pH and clinical dementia rating (A), average neuritic plaque density (B), sum of Consortium to Establish a Registry for Alzheimer’s Disease (CERAD) rating scores in multiple brain regions (C), and sum of neurofibrillary tangles density in multiple brain regions (D). The solid line indicates the regression line, and the dashed line indicates the 95% confidence interval. *r*, Pearson correlation coefficient.

**Supplementary Figure 4. No significant effect of *APOE*_ε_4 status on brain pH.** Forest plot of meta-analysis comparing postmortem brain pH between *APOE*ε4 carriers and non-carriers in control subjects (A; Hedges’ *g* = –0.40, 95% CI = [-0.80; 0.0056], *p* = 0.053) and patients with AD (B; Hedges’ *g* = 0.046, 95% CI = [-0.31; 0.40], *p* = 0.80). 95% CI, 95% confidence interval; SE, standard error; SMD, standardized mean difference.

## References

1. Atwood CS, Moir RD, Huang X, Scarpa RC, Bacarra NME, Romano DM, Hartshorn MA, Tanzi RE, Bush AI (1998) Dramatic Aggregation of Alzheimer Aβ by Cu(II) Is Induced by Conditions Representing Physiological Acidosis. J Biol Chem 273:12817–12826.

2. Basurto-Islas G, Grundke-Iqbal I, Tung YC, Liu F, Iqbal K (2013) Activation of Asparaginyl Endopeptidase Leads to Tau Hyperphosphorylation in Alzheimer Disease. J Biol Chem 288:17495–17507.

3. Belloy ME, Napolioni V, Greicius MD (2019) A Quarter Century of APOE and Alzheimer’s Disease: Progress to Date and the Path Forward. Neuron 101:820–838.

4. Braak H, Braak E (1991) Neuropathological stageing of Alzheimer-related changes. Acta Neuropathol (Berl) 82:239–259.

5. Busche MA, Konnerth A (2016) Impairments of neural circuit function in Alzheimer’s disease. Philos Trans R Soc B Biol Sci 371:20150429.

6. Chesler M, Kaila K (1992) Modulation of pH by neuronal activity. Trends Neurosci 15:396–402.

7. Decker Y, Németh E, Schomburg R, Chemla A, Fülöp L, Menger MD, Liu Y, Fassbender K (2021) Decreased pH in the aging brain and Alzheimer’s disease. Neurobiol Aging 101:40–49.

8. Dogan AE, Yuksel C, Du F, Chouinard V-A, Öngür D (2018) Brain lactate and pH in schizophrenia and bipolar disorder: a systematic review of findings from magnetic resonance studies. Neuropsychopharmacology 43:1681–1690.

9. Hagihara H et al. (2024) Large-scale animal model study uncovers altered brain pH and lactate levels as a transdiagnostic endophenotype of neuropsychiatric disorders involving cognitive impairment. eLife 12:RP89376.

10. Hagihara H, Catts VS, Katayama Y, Shoji H, Takagi T, Huang FL, Nakao A, Mori Y, Huang K-P, Ishii S, Graef IA, Nakayama KI, Shannon Weickert C, Miyakawa T (2018) Decreased brain pH as a shared endophenotype of psychiatric disorders. Neuropsychopharmacology 43:459–468.

11. Hagihara H, Murano T, Miyakawa T (2023) The gene expression patterns as surrogate indices of pH in the brain. Front Psychiatry 14:1151480.

12. Halim ND, Lipska BK, Hyde TM, Deep-Soboslay A, Saylor EM, Herman M, Thakar J, Verma A, Kleinman JE (2008) Increased lactate levels and reduced pH in postmortem brains of schizophrenics: medication confounds. J Neurosci Methods 169:208–213.

13. Hirata K et al. (2024) Altered Brain Energy Metabolism Related to Astrocytes in Alzheimer’s Disease. Ann Neurol 95:104–115.

14. Iaccarino L, Tammewar G, Ayakta N, Baker SL, Bejanin A, Boxer AL, Gorno-Tempini ML, Janabi M, Kramer JH, Lazaris A, Lockhart SN, Miller BL, Miller ZA, O’Neil JP, Ossenkoppele R, Rosen HJ, Schonhaut DR, Jagust WJ, Rabinovici GD (2018) Local and distant relationships between amyloid, tau and neurodegeneration in Alzheimer’s Disease. NeuroImage Clin 17:452–464.

15. Kobayashi S, Tanaka T, Soeda Y, Almeida OFX, Takashima A (2017) Local Somatodendritic Translation and Hyperphosphorylation of Tau Protein Triggered by AMPA and NMDA Receptor Stimulation. eBioMedicine 20:120–126.

16. Kobayashi S, Tanaka T, Soeda Y, Takashima A (2019) Enhanced Tau Protein Translation by Hyper-Excitation. Front Aging Neurosci 11:322.

17. Lewis DA (2002) The Human Brain Revisited: Opportunities and Challenges in Postmortem Studies of Psychiatric Disorders. Neuropsychopharmacology 26:143–154.

18. Liguori C, Chiaravalloti A, Sancesario G, Stefani A, Sancesario GM, Mercuri NB, Schillaci O, Pierantozzi M (2016) Cerebrospinal fluid lactate levels and brain [18F]FDG PET hypometabolism within the default mode network in Alzheimer’s disease. Eur J Nucl Med Mol Imaging 43:2040–2049.

19. Liguori C, Stefani A, Sancesario G, Sancesario GM, Marciani MG, Pierantozzi M (2015) CSF lactate levels, τ proteins, cognitive decline: a dynamic relationship in Alzheimer’s disease. J Neurol Neurosurg Psychiatry 86:655–659.

20. Lyros E, Ragoschke-Schumm A, Kostopoulos P, Sehr A, Backens M, Kalampokini S, Decker Y, Lesmeister M, Liu Y, Reith W, Fassbender K (2020) Normal brain aging and Alzheimer’s disease are associated with lower cerebral pH: an in vivo histidine 1H-MR spectroscopy study. Neurobiol Aging 87:60–69.

21. Maestú F, de Haan W, Busche MA, DeFelipe J (2021) Neuronal excitation/inhibition imbalance: core element of a translational perspective on Alzheimer pathophysiology. Ageing Res Rev 69:101372.

22. Mandal PK, Akolkar H, Tripathi M (2012) Mapping of hippocampal pH and neurochemicals from in vivo multi-voxel 31P study in healthy normal young male/female, mild cognitive impairment, and Alzheimer’s disease. J Alzheimers Dis JAD 31 Suppl 3:S75–86.

23. Mecheri G, Marie-Cardine M, Sappey-Marinier D, Bonmartin H, Albrand G, Ferry G, Coppard-Meyer N, Courpron P (1997) In vivo hippocampal (31)P NMR metabolites in Alzheimer’s disease and ageing. Eur Psychiatry J Assoc Eur Psychiatr 12:140–148.

24. Montine TJ, Phelps CH, Beach TG, Bigio EH, Cairns NJ, Dickson DW, Duyckaerts C, Frosch MP, Masliah E, Mirra SS, Nelson PT, Schneider JA, Thal DR, Trojanowski JQ, Vinters HV, Hyman BT (2012) National Institute on Aging–Alzheimer’s Association guidelines for the neuropathologic assessment of Alzheimer’s disease: a practical approach. Acta Neuropathol (Berl) 123:1–11.

25. Mullins R, Reiter D, Kapogiannis D (2018) Magnetic resonance spectroscopy reveals abnormalities of glucose metabolism in the Alzheimer’s brain. Ann Clin Transl Neurol 5:262–272.

26. Murano T, Hagihara H, Tajinda K, Matsumoto M, Miyakawa T (2019) Transcriptomic immaturity inducible by neural hyperexcitation is shared by multiple neuropsychiatric disorders. Commun Biol 2:32.

27. Page MJ et al. (2021) The PRISMA 2020 statement: An updated guideline for reporting systematic reviews. Int J Surg 88:105906.

28. Palop JJ, Chin J, Roberson ED, Wang J, Thwin MT, Bien-Ly N, Yoo J, Ho KO, Yu G-Q, Kreitzer A, Finkbeiner S, Noebels JL, Mucke L (2007) Aberrant excitatory neuronal activity and compensatory remodeling of inhibitory hippocampal circuits in mouse models of Alzheimer’s disease. Neuron 55:697–711.

29. Pavlov I, Kaila K, Kullmann DM, Miles R (2013) Cortical inhibition, pH and cell excitability in epilepsy: what are optimal targets for antiepileptic interventions?: Cortical inhibition, pH and cell excitability in epilepsy. J Physiol 591:765–774.

30. Pellerin L, Magistretti PJ (1994) Glutamate uptake into astrocytes stimulates aerobic glycolysis: a mechanism coupling neuronal activity to glucose utilization. Proc Natl Acad Sci 91:10625–10629.

31. Prabakaran S, Swatton JE, Ryan MM, Huffaker SJ, Huang JT-J, Griffin JL, Wayland M, Freeman T, Dudbridge F, Lilley KS, Karp NA, Hester S, Tkachev D, Mimmack ML, Yolken RH, Webster MJ, Torrey EF, Bahn S (2004) Mitochondrial dysfunction in schizophrenia: evidence for compromised brain metabolism and oxidative stress. Mol Psychiatry 9:684–697.

32. Prince M, Wimo A, Guerchet M, Ali G-C, Wu Y-T, Prina M (2015) World Alzheimer Report 2015. The Global Impact of Dementia: An analysis of prevalence, incidence, cost and trends. Alzheimer’s Dis Int hal-03495438.

33. Pruett BS, Meador-Woodruff JH (2020) Evidence for altered energy metabolism, increased lactate, and decreased pH in schizophrenia brain: A focused review and meta-analysis of human postmortem and magnetic resonance spectroscopy studies. Schizophr Res 223:29–42.

34. Rijpma A, van der Graaf M, Meulenbroek O, Olde Rikkert MGM, Heerschap A (2018) Altered brain high-energy phosphate metabolism in mild Alzheimer’s disease: A 3-dimensional 31P MR spectroscopic imaging study. NeuroImage Clin 18:254–261.

35. Saunders AM, Roses AD, Pericak-Vance MA, Dole KC, Strittmatter WJ, Schmechel DE, Szymanski MH, McCown N, Manwaring MG, Schmader K, Breitner JCS, Goldgaber D, Benson MD, Goldfarb L, Brown WT (1993) Apolipoprotein E ∈4 allele distributions in late-onset Alzheimer’s disease and in other amyloid-forming diseases. The Lancet 342:710–711.

36. Scheltens P, Blennow K, Breteler MMB, de Strooper B, Frisoni GB, Salloway S, Van der Flier WM (2016) Alzheimer’s disease. The Lancet 388:505–517.

37. Schuchmann S, Schmitz D, Rivera C, Vanhatalo S, Salmen B, Mackie K, Sipilä ST, Voipio J, Kaila K (2006) Experimental febrile seizures are precipitated by a hyperthermia-induced respiratory alkalosis. Nat Med 12:817–823.

38. Serrano-Pozo A, Das S, Hyman BT (2021) APOE and Alzheimer’s disease: advances in genetics, pathophysiology, and therapeutic approaches. Lancet Neurol 20:68–80.

39. Siesjö BK (1985) Acid-Base Homeostasis in the Brain: Physiology, Chemistry, and Neurochemical Pathology. In: Progress in Brain Research (K. Kogure K-AH BK Siesjö and FA Welsh, ed), pp 121–154 Molecular Mechanisms of Ischemic Brain Damage. Elsevier.

40. Sims R, Hill M, Williams J (2020) The multiplex model of the genetics of Alzheimer’s disease. Nat Neurosci 23:311–322.

41. Stork C, Renshaw PF (2005) Mitochondrial dysfunction in bipolar disorder: evidence from magnetic resonance spectroscopy research. Mol Psychiatry 10:900–919.

42. Tomita H, Vawter MP, Walsh DM, Evans SJ, Choudary PV, Li J, Overman KM, Atz ME, Myers RM, Jones EG, Watson SJ, Akil H, Bunney Jr. WE (2004) Effect of agonal and postmortem factors on gene expression profile: quality control in microarray analyses of postmortem human brain. Biol Psychiatry 55:346–352.

43. Wilkosz PA, Seltman HJ, Devlin B, Weamer EA, Lopez OL, DeKosky ST, Sweet RA (2010) Trajectories of cognitive decline in Alzheimer’s disease. Int Psychogeriatr 22:281–290.

44. You JC, Muralidharan K, Park JW, Petrof I, Pyfer MS, Corbett BF, LaFrancois JJ, Zheng Y, Zhang X, Mohila CA, Yoshor D, Rissman RA, Nestler EJ, Scharfman HE, Chin J (2017) Epigenetic suppression of hippocampal calbindin-D28k by ΔFosB drives seizure-related cognitive deficits. Nat Med 23:1377–1383.

45. Zheng W-H, Bastianetto S, Mennicken F, Ma W, Kar S (2002) Amyloid β peptide induces tau phosphorylation and loss of cholinergic neurons in rat primary septal cultures. Neuroscience 115:201–211.

