## Supplementary Figures 1-4 for "Decreased brain pH correlated with progression of Alzheimer’s disease neuropathology: a systematic review and meta-analyses of postmortem studies"

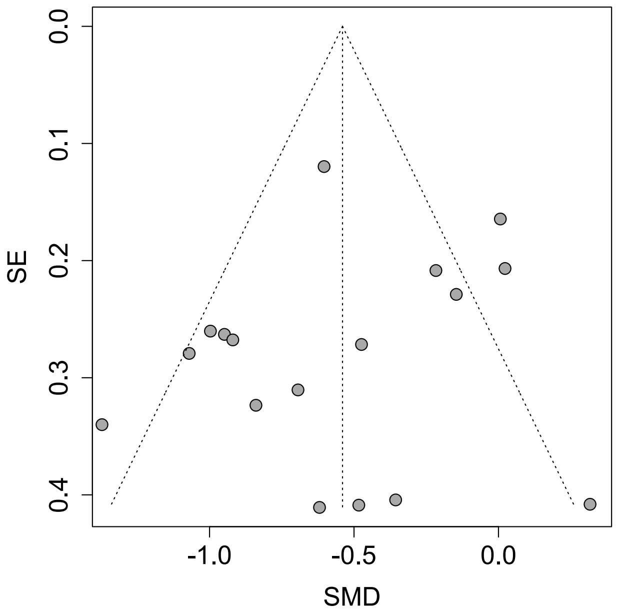


**Supplementary Figure 1. Funnel plot of the included datasets.** Each plot represents a dataset. SE, standard error; SMD, standardized mean difference.

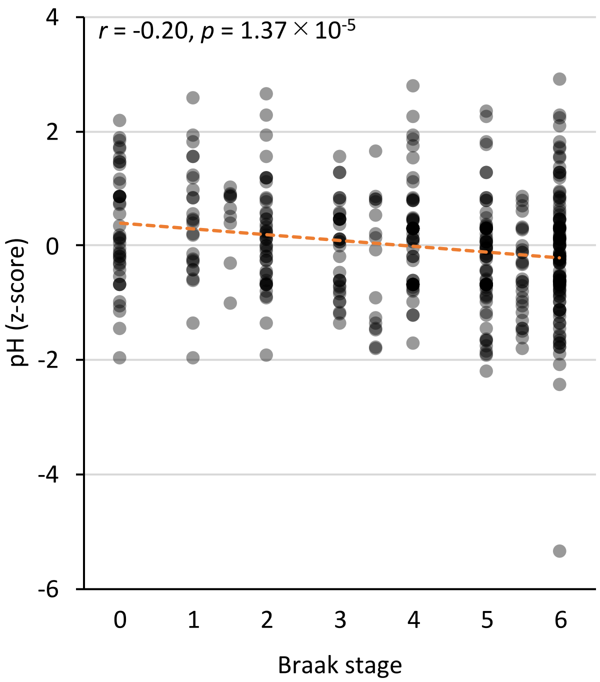


**Supplementary Figure 2. Scatter plot depicting the negative correlation between pH (z-score) and Braak stages.** A z-score was calculated for each subject within the dataset to standardize pH measurements. Each dot represents the data for an individual subject. r, Pearson correlation coefficient.

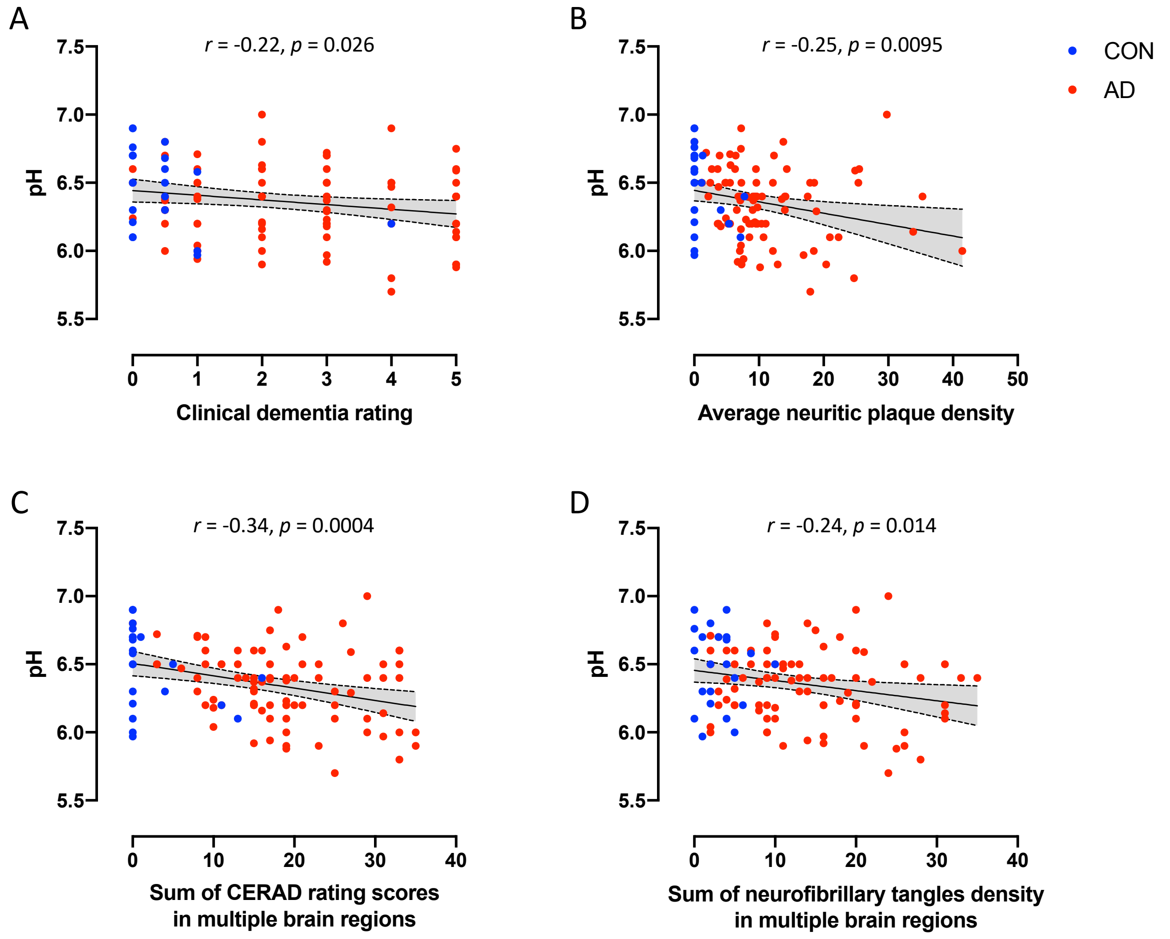


**Supplementary Figure 3. Significant correlation between a decrease in brain pH and an increase in disease severity.** Data of GSE84422 was analyzed. Scatter plot of correlation between brain pH and clinical dementia rating (A), average neuritic plaque density (B), sum of Consortium to Establish a Registry for Alzheimer's Disease (CERAD) rating scores in multiple brain regions (C), and sum of neurofibrillary tangles density in multiple brain regions (D). The solid line indicates the regression line, and the dashed line indicates the 95% confidence interval. *r*, Pearson correlation coefficient.

A

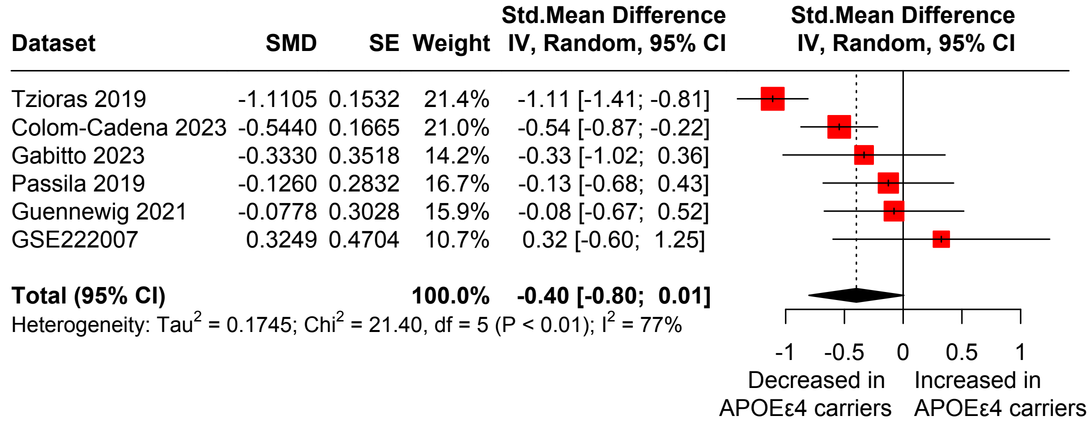


B

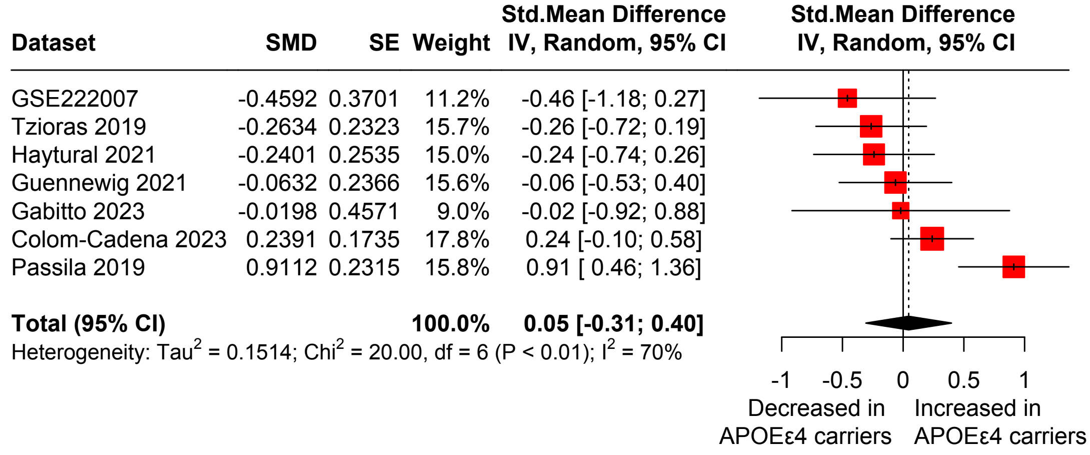


**Supplementary Figure 4. No significant effect of *APOE*ε4 status on brain pH.** Forest plot of meta-analysis comparing postmortem brain pH between *APOE*ε4 carriers and non-carriers in control subjects (A; Hedges' $g$ = -0.40, 95% CI = [-0.80; 0.0056], $p$ = 0.053) and patients with AD (B; Hedges' $g$ = 0.046, 95% CI = [-0.31; 0.40], $p$ = 0.80). 95% CI, 95% confidence interval; SE, standard error; SMD, standardized mean difference.
