## Supplementary Table 1 for "Decreased brain pH correlated with progression of Alzheimer’s disease neuropathology: a systematic review and meta-analyses of postmortem studies"

**Supplementary Table 1.** Summary of datasets used in this study

| Dataset name | Diagnosis /group | Number of samples (Female/Male) | Age (years)$^{\#}$ | PMI (hours)$^{\#}$ | pH$^{\#}$ | Sample | Source/ Reference | Data analysis | | |
|---|---|---|---|---|---|---|---|---|---|---|
| | | | | | | | | Fig. 2 | Fig. 3 | Fig. 4 |
| Barrachina 2006 | CON<br>AD | 6 (3/3)<br>16 (7/9) | 71.3 ± 3.4<br>76.8 ± 2.0 | 8.0 ± 1.2<br>5.1 ± 0.6$^{*}$ | 6.58 ± 0.11<br>6.54 ± 0.05 | Brain (cerebral cortex) | [1] | y | y | n |
| Bayer 2009, 2014, Coulson 2010, Quinn 2012 | CON<br>AD | 5 (3/2)<br>28 (19/9) | 78.2 ± 3.5<br>80.3 ± 2.1 | 5.7 ± 0.4<br>5.7 ± 0.9 | 6.58 ± 0.05<br>6.54 ± 0.04 | CSF | [2–5] | y | y | n |
| Colom-Cadena 2023 | CON<br>AD | 19 (7/12)<br>24 (12/12) | 81.0 ± 0.6<br>83.3 ± 2.0 | 64.0 ± 6.1<br>76.5 ± 5.0 | 6.08 ± 0.04<br>6.08 ± 0.03 | Brain | [6] | y | y | y |
| Ervin 2007 | CON<br>AD | 36 (19/17)<br>32 (24/8) | 82.4 ± 1.9<br>83.6 ± 0.6 | 10.5 ± 1.3<br>7.6 ± 0.9 | 7.06 ± 0.15<br>6.85 ± 0.09 | Postmortem ventricular fluid | [7] | y | n | n |
| Gabitto 2023 | CON<br>AD | 42 (24/18)<br>42 (27/15) | 82.8 ± 1.0<br>80.7 ± 1.8 | 6.9 ± 0.4<br>7.1 ± 0.3 | 6.75 ± 0.05<br>6.60 ± 0.07 | Brain | [8] | y | y | y |
| Garamszegi 2023 | CON<br>AD | 1 (NA)<br>5 (NA) | NA<br>NA | 26.5<br>11.0 ± 2.8 | 6.23<br>5.94 ± 0.13 | Brain | [9] | n | y | n |
| Griswold 2021 | CON<br>AD | 0<br>11 (9/2) | –<br>NA | –<br>9.4 ± 2.3 | –<br>6.34 ± 0.06 | Brain (frontal cortex) | [10] | n | y | n |
| GSE222007 | CON<br>AD | 10 (5/5)<br>10 (5/5) | 88.3 ± 1.2<br>88.4 ± 1.3 | 42.3 ± 7.1<br>38.2 ± 5.8 | 5.87 ± 0.14<br>6.00 ± 0.12 | Brain | NCBI GEO | y | y | y |
| GSE44771 | CON<br>AD | 101 (19/82)<br>129 (67/62)$^{\dagger\dagger}$ | 62.1 ± 1.1<br>80.1 ± 0.8$^{**}$ | 22.4 ± 0.6<br>13.7 ± 0.7$^{**}$ | 6.50 ± 0.03<br>6.28 ± 0.03$^{**}$ | Brain | NCBI GEO | y | n | n |
| GSE84422 | CON<br>AD | 22 (14/8)<br>85 (63/22) | 80.7 ± 2.5<br>87.5 ± 0.9$^{**}$ | 7.8 ± 1.4<br>5.7 ± 0.6 | 6.47 ± 0.06<br>6.34 ± 0.03$^{*}$ | Brain | NCBI GEO | y | y | n |
| GSE146639 | CON<br>AD | 14 (8/6)<br>11 (3/8) | 83.7 ± 3.6<br>81.8 ± 3.4 | 3.4 ± 0.9<br>1.7 ± 0.7 | 6.82 ± 0.14<br>6.57 ± 0.07 | CSF | NCBI GEO | y | n | n |
| Guennewig 2021 | CON<br>AD | 22 (12/10)<br>26 (15/11) | 77.2 ± 2.4<br>77.9 ± 2.3 | 22.1 ± 3.5<br>13.0 ± 1.8$^{*}$ | 6.55 ± 0.06<br>6.29 ± 0.05$^{**}$ | Brain (occipital lobe) | [11] | y | n | y |
| Haytural 2021 | CON<br>AD | 7 (7/0)<br>8 (8/0) | 80.4 ± 1.6<br>80.3 ± 3.2 | 6.1 ± 0.3<br>5.1 ± 0.3 | 6.82 ± 0.18<br>6.30 ± 0.08$^{*}$ | Brain | [12] | y | y | y |
| Jacobs 2006 | CON<br>AD | 19 (8/11)<br>32 (15/17) | 74.1 ± 2.9<br>81.2 ± 1.2$^{*}$ | 45.0 ± 6.0<br>38.4 ± 4.4 | 6.58 ± 0.09<br>6.30 ± 0.07$^{*}$ | Brain (cerebellum) | [13] | y | y | n |
| Passila 2019, 2020 | CON<br>AD | 15 (11/4)<br>8 (6/2) | 83.9 ± 2.1<br>83.3 ± 2.4 | 16.2 ± 2.9<br>14.5 ± 4.0 | 6.43 ± 0.07<br>6.17 ± 0.08$^{*}$ | Brain (cerebellum) | [14, 15] | y | y | y |
| Passila 2021 | CON<br>AD | 5 (3/2)<br>7 (5/2) | 74.8 ± 5.8<br>85.3 ± 3.1 | 23.8 ± 5.6<br>9.1 ± 3.0$^{*}$ | 6.53 ± 0.08<br>6.23 ± 0.12 | Brain | [16] | y | y | n |
| Tzioras 2019 | CON<br>AD | 17 (5/12)<br>23 (9/14) | 72.0 ± 2.3<br>77.6 ± 2.4 | 66.2 ± 4.6<br>73.9 ± 4.2 | 6.13 ± 0.04<br>6.13 ± 0.05 | Brain | [17] | y | y | y |
| Ueberham 2006 | CON<br>AD | 5 (NA)<br>5 (NA) | 69.4 ± 2.7<br>75.2 ± 3.0 | 39.2 ± 4.8<br>26.2 ± 3.9 | 6.56 ± 0.06<br>6.48 ± 0.04 | Brain (hippocampus) | [18] | y | y | n |
| Wong 2012 | CON<br>AD | 6 (3/3)<br>6 (3/3) | 81.0 ± 3.3<br>79.2 ± 2.6 | 16.7 ± 2.7<br>9.5 ± 3.7 | 6.58 ± 0.11<br>6.32 ± 0.11 | Brain | [19] | y | y | n |

AD, Alzheimer's disease; CON, non-AD controls; CSF, cerebrospinal fluid; NA, not available; NCBI GEO, National Center for Biotechnology Information Gene Expression Omnibus database; PMI, postmortem interval.

#mean ± sem; †$p < 0.05$, ††$p < 0.01$, chi-square test for the sex ratio; *$p < 0.05$, **$p < 0.01$, two-tailed unpaired t-test.

**Supplementary references**


1. Barrachina M, Castaño E, Ferrer I. TaqMan PCR assay in the control of RNA normalization in human post-mortem brain tissue. Neurochem Int. 2006;49:276–284.

2. Beyer N, Coulson DTR, Quinn JG, Brockbank S, Hellemans J, Irvine GB, et al. mRNA levels of BACE1 and its interacting proteins, RTN3 and PPIL2, correlate in human post mortem brain tissue. Neuroscience. 2014;274:44–52.

3. Beyer N, Coulson DT, Heggarty S, Ravid R, Irvine GB, Hellemans J, et al. ZnT3 mRNA levels are reduced in Alzheimer's disease post-mortem brain. Mol Neurodegener. 2009;4:53.

4. Coulson DTR, Beyer N, Quinn JG, Brockbank S, Hellemans J, Irvine GB, et al. BACE1 mRNA Expression in Alzheimer's Disease Postmortem Brain Tissue. J Alzheimers Dis. 2010;22:1111–1122.

5. Quinn JG, Coulson DTR, Brockbank S, Beyer N, Ravid R, Hellemans J, et al. α-Synuclein mRNA and soluble α-synuclein protein levels in post-mortem brain from patients with Parkinson's disease, dementia with Lewy bodies, and Alzheimer's disease. Brain Res. 2012;1459:71–80.

6. Colom-Cadena M, Davies C, Sirisi S, Lee J-E, Simzer EM, Tzioras M, et al. Synaptic oligomeric tau in Alzheimer's disease — A potential culprit in the spread of tau pathology through the brain. Neuron. 2023;111:2170-2183.e6.

7. Ervin JF, Heinzen EL, Cronin KD, Goldstein D, Szymanski MH, Burke JR, et al. Postmortem Delay Has Minimal Effect on Brain RNA Integrity. J Neuropathol Exp Neurol. 2007;66:1093–1099.

8. Gabitto M. Integrated multimodal cell atlas of Alzheimer's disease. 2023. https://www.researchsquare.com. Accessed 12 October 2023.

9. Garamszegi SP, Banack SA, Duque LL, Metcalf JS, Stommel EW, Cox PA, et al. Detection of β-N-methylamino-l-alanine in postmortem olfactory bulbs of Alzheimer's disease patients using UHPLC-MS/MS: An autopsy case-series study. Toxicol Rep. 2023;10:87–96.

10. Griswold AJ, Celis K, Bussies PL, Rajabli F, Whitehead PL, Hamilton-Nelson KL, et al. Increased APOE ε4 expression is associated with the difference in Alzheimer's disease risk from diverse ancestral backgrounds. Alzheimers Dement. 2021;17:1179–1188.

11. Guennewig B, Lim J, Marshall L, McCorkindale AN, Paasila PJ, Patrick E, et al. Defining early changes in Alzheimer's disease from RNA sequencing of brain regions differentially affected by pathology. Sci Rep. 2021;11:4865.

12. Haytural H, Jordà-Siquier T, Winblad B, Mulle C, Tjernberg LO, Granholm A-C, et al. Distinctive alteration of presynaptic proteins in the outer molecular layer of the dentate gyrus in Alzheimer's disease. Brain Commun. 2021;3:fcab079.

13. Jacobs EH, Williams RJ, Francis PT. Cyclin-dependent kinase 5, Munc18a and Munc18-interacting protein 1/X11α protein up-regulation in Alzheimer's disease. Neuroscience. 2006;138:511–522.

14. Paasila PJ, Davies DS, Kril JJ, Goldsbury C, Sutherland GT. The relationship between the morphological subtypes of microglia and Alzheimer's disease neuropathology. Brain Pathol. 2019;29:726–740.

15. Paasila PJ, Davies DS, Sutherland GT, Goldsbury C. Clustering of activated microglia occurs before the formation of dystrophic neurites in the evolution of Aβ plaques in Alzheimer's disease. Free Neuropathol. 2020;1:1–20.

16. Paasila PJ, Fok SYY, Flores-Rodriguez N, Sajjan S, Svahn AJ, Dennis CV, et al. Ground state depletion microscopy as a tool for studying microglia–synapse interactions. J Neurosci Res. 2021;99:1515–1532.

17. Tzioras M, Daniels MJD, King D, Popovic K, Holloway RK, Stevenson AJ, et al. Altered synaptic ingestion by human microglia in Alzheimer's disease. 2019:795930.

18. Ueberham U, Ueberham E, Gruschka H, Arendt T. Altered subcellular location of phosphorylated Smads in Alzheimer's disease. Eur J Neurosci. 2006;24:2327–2334.

19. Wong J, Garner B, Halliday GM, Kwok JBJ. Srp20 regulates TrkB pre-mRNA splicing to generate TrkB-Shc transcripts with implications for Alzheimer's disease. J Neurochem. 2012;123:159–171.
